# DOC screen completion time reflects executive function, speed of processing and fluency in an observational cohort study

**DOI:** 10.1101/2023.02.15.23286004

**Authors:** Alisia Southwell, Sajeevan Sujanthan, Tera Armel, Elaine Xing, Arunima Kapoor, Xiao Yu Eileen Liu, Krista L. Lanctot, Nathan Herrmann, Brian J. Murray, Kevin E. Thorpe, Megan L. Cayley, Michelle N. Sicard, Karen Lien, Demetrios J. Sahlas, Richard H. Swartz

## Abstract

**Background:** The DOC screen was developed to identify Depression, Obstructive sleep apnea, and Cognitive impairment (“DOC” comorbidities) after stroke. Each component has its own score, but additional information may be gained from the time to complete the screen. Cognitive screening completion time is rarely used as an outcome measure. We assessed the added value of using DOC screen completion time as a predictor of impairment on detailed cognitive assessments.

**Methods:** Consecutive English-speaking new referrals to the stroke prevention clinic were consented to participate in detailed neuropsychological testing (n=437). DOC screen scores and times were compared to cognitive test scores using multiple linear regression and receiver operating characteristic (ROC) analysis. All linear regression analyses controlled for age, sex, years of education, and functional outcome as assessed by the modified Rankin score.

**Results:** Average completion time for the DOC screen was 3.8 ± 1.3 minutes. After accounting for age, sex and cognitive screen score, completion time was a significant independent predictor, of speed of processing (p = .002, 95% CI: -0.016 to -0.004), verbal fluency (p < .001, CI: -0.012 to -0.006) and executive (p = .004, CI: -0.006 to -0.001), but not memory, function. Completion time above 5.5 minutes (332.5 seconds) was associated with a high likelihood of impairment on gold standard executive (likelihood ratios 3.9-5.2) and speed of processing (likelihood ratio = 5.2) tasks.

**Conclusions:** DOC screen completion time is easy to collect in routine care and is independently associated with speed of processing, language and executive dysfunctions after stroke. People who take more than 5.5 minutes to complete the DOC screen are likely to have deficits in executive functioning and speed of processing. These domains can be challenging to screen for in stroke survivors, and this measure provides a simple, clinically feasible method to screen for these under-appreciated concerns.

Clinical Trials Registration Identifier: NCT02363114

Clinical Trials URL: https://clinicaltrials.gov/ct2/show/NCT02363114

## Introduction

Stroke is the leading cause of neurological disability in adults^1^ and survival after stroke is increasing.^2–4^ In addition to physical post-stroke deficits,^5^ approximately 30 to 50% of stroke survivors are affected by each of **d**epression, **o**bstructive sleep apnea (OSA), and **c**ognitive impairment (DOC).^6–9^ These DOC comorbidities are all associated with poorer functional outcomes,^10^ and an increased risk of mortality.^11^

The DOC screen was developed as a feasible and valid tool to screen and stratify stroke patients into high, intermediate, and low risk groups for DOC comorbidities to facilitate detection and management in high-volume stroke clinic settings.^12^ The screen is efficient, yet designed to maintain the construct validity of a delayed recall task. Eighty-nine percent of patients in stroke prevention clinics are able to complete the tool in <6 minutes (mean=4.2 minutes, SD=1.5).^12^ In validation studies, the cognitive component of the DOC score is helpful to quickly stratify people into “cognitively normal”, “cognitively impaired” and “need more assessment” groups, compared to more detailed cognitive testing.^12^Although the DOC completion time was originally collected as a way to assess feasibility, practitioners can record this measure when administering the DOC screen in clinical settings. Several studies have reported the average time taken to complete other well-known cognitive screens as feasibility demonstrations, including the Montreal Cognitive Assessment (MoCA; means ranging from 9.5 minutes – 11 minutes)^13,14^ and the Mini Mental State Examination (MMSE; means ranging from 8 minutes – 13.4 minutes).^14,15^ However, few studies have assessed the utility of using a cognitive screen’s completion time as a metric to evaluate underlying cognitive abilities, such as executive functioning.

Executive dysfunction and delays in speed of processing are the most commonly reported cognitive impairments after stroke. The DOC screen specifically examines mood symptoms, cognitive (executive, memory and abstraction) dysfunction and OSA/fatigue – all of which could be associated with cognitive or psychomotor slowing.^16^ Screen completion time is an immediately available metric, requiring no additional effort from either patients or clinicians, that might reflect executive function. The objective of this study was to determine whether completion time for the DOC screen is a reliable reflection of cognitive dysfunction and whether a single completion time cut-point could indicate impairment.

## Methods

All patients were recruited from the DOC feasibility and validity study.^12^ This study included English speaking (or English fluent) patients newly referred to stroke prevention clinics over a two-year period (n=1504). We excluded patients with severe aphasia, severe motor dysfunction and patients who were not fluent in English. Each eligible participant was administered the DOC screen (**Figure 1**) as a brief screen of depression, obstructive sleep apnea (OSA) and cognitive impairment. All DOC screens were timed from the beginning of the memory registration (first task) until the end of the 5-word free recall (final task). Chart abstractions by trained research members captured demographic and clinical data on all participants from patient charts using previously published and validated methods.^17,18^

**Figure 1:**
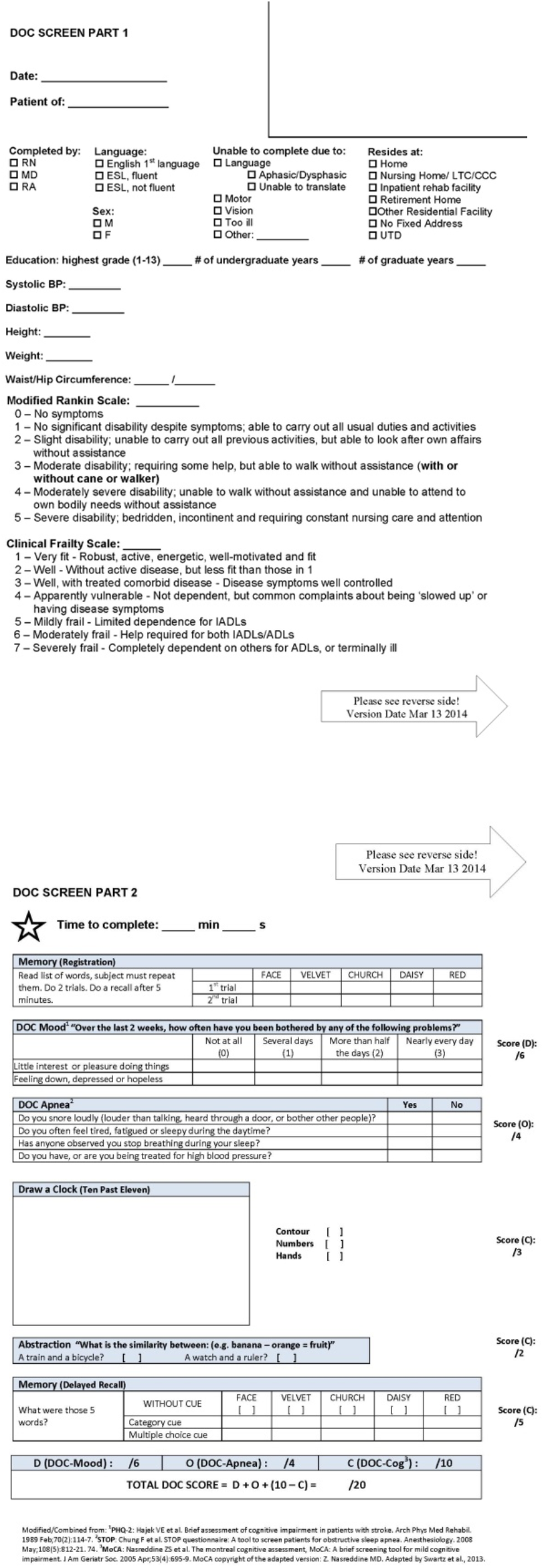
The DOC screen (freely available for download at www.docscreen.ca).

To reduce sampling bias, all consecutive patients from stroke prevention clinics who completed the DOC screen were asked to complete more detailed neuropsychological assessments, including a cognitive battery and formal mood assessments as outlined in the DOC feasibly study.^12^ A complete list of all mood and cognitive assessments completed as part of the DOC study is reported elsewhere.^12^ In this analysis, cognition was assessed using the 30-minute neuropsychological battery recommended by the NINDS-CSN.^19^ This cognitive battery consists of the: Controlled Oral World Association Test (phonemic fluency), Animal Naming task (semantic fluency), California Verbal Learning Test (CVLT), Digit Symbol Coding, and Trail Making Tests (TMT-A and TMT-B). All scores were normalized (z-score or scaled score) for age using age-matched norms from each respective test manual. CVLT and Animal Naming were also education-standardized.^20,21^ The study was approved by the institutional Research Ethics Board.

### Statistical Analysis

Statistical analysis was performed using IBM SPSS Statistics for Windows version 24. Descriptive statistics, including means and standard deviations, were calculated for age, screen completion time, and number of years of education. To assess whether screen time reflects cognitive function, independent linear regression models were used to examine the association between DOC completion time and the scaled or z-scores of all neuropsychological subtests. Data from all participants was used in the regression models. A sensitivity analysis was performed using a complete case approach to assess whether missing variables affected the models. All models controlled for age, education, modified Rankin Score (mRS) and sex. Due to the established relationship between the DOC cognitive sub-scores and detailed cognitive assessments,^12^ we also controlled for the DOC-Cognition score in all models. Significance was set at p < 0.05 for all analyses. To identify whether a single cut point (in seconds) for screen time could be found with high specificity and likelihood ratios for cognitive impairment, receiver operating characteristic (ROC) curves were used. ROC analyses were run for each neuropsychological assessment significantly associated with the DOC screen completion time. A logistic regression with screen time completion and the impairment classification on the NINDS-CSN assessments was applied to the ROC curves. The classification of impairment of NINDS-CSN was defined as scores >2.0 SD from expected norms, on 2 or more cognitive tasks. This required participants to have completed all tests in the detailed cognitive battery, thus a complete case approach was used for all ROC analyses. First, a single, specific cut-point (time in seconds) was defined based on the ROC curve output for patients with an overall classification of impaired on the NINDS-CSN battery. The cut-point was pre-specified to have 95% specificity for impairment. This cut-point was then applied to ROC curves from each individual assessment and evaluated using likelihood ratios (LR).

## Results

437 patients completed cognitive and mood gold standard assessments (Supplemental Table 1). 213 (48.7%) participants were male, the mean (± standard deviation) age was 62.7 ± 15.6 years, and the mean years of education was 15.6 ± 3.9 years (**Table 1**). The DOC screen completion mean was 3.8 ± 1.3 minutes (range: 1.9-9.6 minutes). One hundred and thirty-four (31%) patients had an ischemic stroke, 138 (32%) had a probable/possible TIA, and the remainder (37%) were diagnosed with other conditions (**Table 1**). Non-stroke/TIA diagnoses included patients referred with possible stroke symptoms, but whose further investigations revealed alternative diagnoses, as well as patients without specific stroke/TIA symptoms referred for either vascular risk reduction or assessment of incidental abnormal imaging findings.

**Table 1:** Demographics for participants completing detailed cognitive and neuropsychological assessments (n = 437).

| Variables | Mean (SD) |
| --- | --- |
| Age (years) | 62.7 (15.6) |
| Education (years) | 15.6 (3.9) |
| DOC screen completion time (s) | 227.8 (76.6) |
| Language | **n (%)** |
| English | 363 (83.1) |
| ESL | 74 (16.9) |
| Sex (female) | 51.3% |
| Most Responsible Diagnosis |  |
| Undetermined Diagnosis | 4 (.9) |
| Abnormal CT/MRI Scan | 21 (4.8) |
| Asymptomatic Carotid Artery Disease | 4 (0.9) |
| Definite Ischemic Stroke | 121 (27.7) |
| Definite TIA | 54 (12.4) |
| Hemorrhage ICH | 17 (3.9) |
| Hemorrhage IVH | 1 (0.2) |
| Hemorrhage SAH | 4 (0.9) |
| Hemorrhage SDH | 1 (0.2) |
| Other Non-Vascular | 96 (22) |
| Other Vascular | 14 (3.2) |
| Possible/Query Ischemic | 13 (3.0) |
| Possible/Query TIA | 84 (19.2) |
| Sinovenous Thrombosis | 3 (0.7) |
| Modified Ranking Scale (mRS) |  |
| 0 | 230 (52.6) |
| 1 | 113 (25.9) |
| 2 | 69(15.8) |
| 3 | 19 (4.3) |
| 4 | 2 (.5) |
| Missing | 4 (.9) |

† TIA = transient ischemic attack, ICH = intracerebral hemorrhage, IVH = intraventricular hemorrhage, SAH = subarachnoid hemorrhage, SDH = subdural hemorrhage

We performed linear regressions with DOC screen completion time (seconds) as a predictor for each neuropsychological assessment score (**Table 2**). In all models we controlled for age, sex, years of education, screening score of cognitive function (DOC-Cognition score), and overall function (mRS). All regression models for screen completion time were significant (p < .001) (Supplemental Table 2). Additionally, model summaries showed that screen completion time was a significant predictor (p < 0.005) of: verbal fluency semantic score (95% Confidence Interval (CI) of Beta-coefficient from linear regression: -.006 to -.001), verbal fluency phonemic Score (95% CI: -.018 to -.006), Digit Symbol Coding (95% CI: -.016 to -.004) and the Trail Making Tests (TMT-A 95% CI: -.017 to -.005; TMT-B 95% CI: -.016 to -.004). In all cases, these were negative correlations (i.e., longer completion times correlated with poorer cognitive scores). DOC screen completion time was not a significant predictor of memory performance on the CVLT Short Delay Free Recall (p=.713, 95% CI: -.003 to .002) or the CVLT Long Delay Free Recall (p=.790, 95% CI: -.002 to .003). Results did not differ in the sensitivity models requiring complete case data (see Table 2 compared to Supplemental Table 3 with complete case data).

**Table 2:**
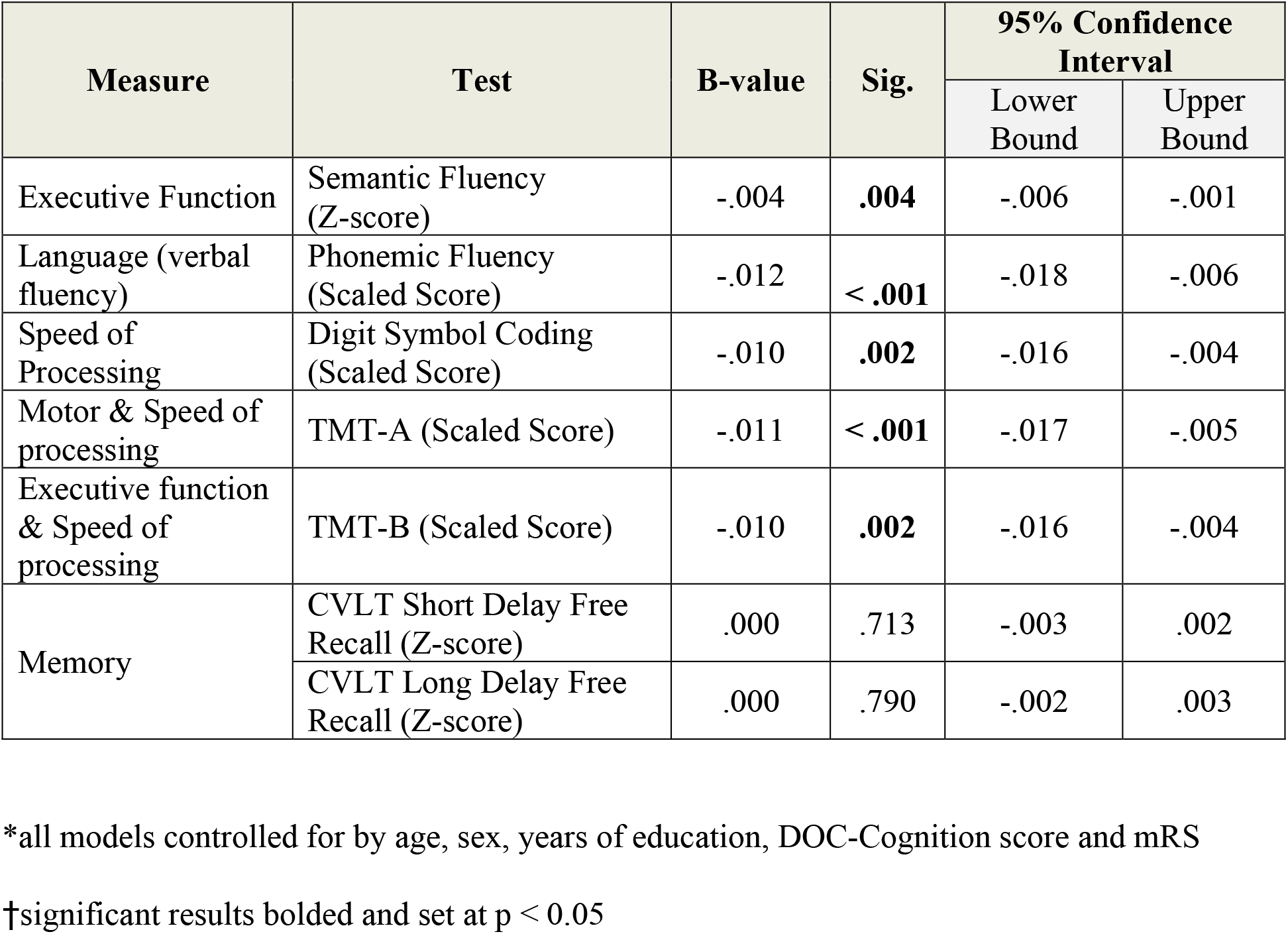
Linear Regression results showing the effect of DOC screen completion time on individual neuropsychological assessments.

Using the single cut-off point approach on the overall impairment ROC curve (Figure 2, Table 3A), the point with 95% specificity for impairment was 332.5 seconds. When this time was applied to ROC models for each individual cognitive task (Table 3B), the same cut-point had high specificity on all executive and speed of processing tasks. The area under the curve was greater than 0.7 for all executive and speed of processing tasks. Likelihood ratios for predicting abnormal results on executive and speed of processing tasks ranged from four to six – that is, people taking more than 332.5 seconds to complete the DOC screen were 4-6 times more likely to have severe impairment on executive and speed of processing tasks than those with faster completion times (see Table 3).

**Table 3:**
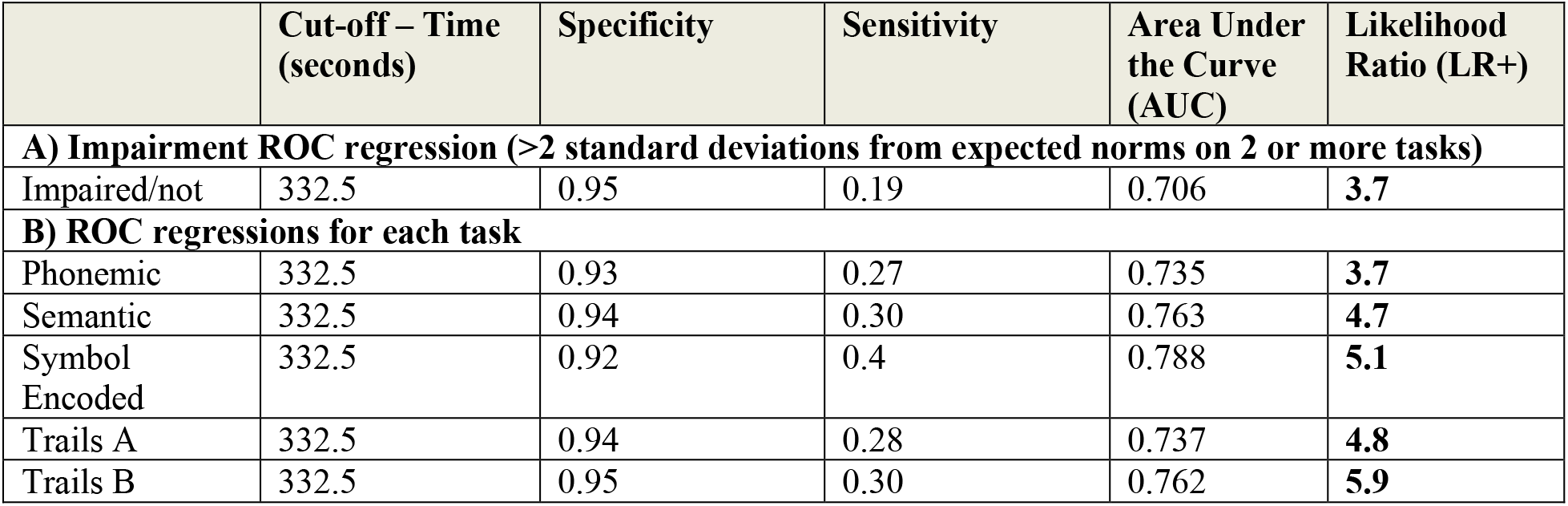
– ROC model outputs comparing DOC screen completion time with full neuropsychological assessments, with a cut off set at **332.5 seconds (95% specificity) obtained from the model for overall impairment**.

|  | Cut-off – Time<br>(seconds) | Specificity | Sensitivity | Area Under<br>the Curve<br>(AUC) | Likelihood<br>Ratio (LR+) |
| --- | --- | --- | --- | --- | --- |
| **A) Impairment ROC regression (>2 standard deviations from expected norms on 2 or more tasks)** |  |  |  |  |  |
| Impaired/not | 332.5 | 0.95 | 0.19 | 0.706 | **3.7** |
| **B) ROC regressions for each task** |  |  |  |  |  |
| Phonemic | 332.5 | 0.93 | 0.27 | 0.735 | **3.7** |
| Semantic | 332.5 | 0.94 | 0.30 | 0.763 | **4.7** |
| Symbol<br>Encoded | 332.5 | 0.92 | 0.4 | 0.788 | **5.1** |
| Trails A | 332.5 | 0.94 | 0.28 | 0.737 | **4.8** |
| Trails B | 332.5 | 0.95 | 0.30 | 0.762 | **5.9** |

## Discussion

Several studies^22^ have shown that post-stroke impairments can be separated into independent cognitive factors including language, memory and executive function, with deficits in executive functioning and speed of processing being the most common.^23^ Screening tests for executive function and speed of processing are limited in routine clinical care. These results demonstrate that DOC screen completion time is an independent predictor of executive function (semantic fluency,^24^ TMT-B^25^), speed of processing (Digit symbol coding,^26^ TMT-A and B^27^) and verbal fluency^28^ after stroke, even after controlling for age, sex, education, DOC-cognition score and stroke severity. Completion time did not predict CVLT scores, a verbal test primarily affecting verbal memory (learning/registration and recall).^29^ Verbal fluency, while reflecting language function, is also reflective of executive function.^30^ Moreover, we have demonstrated that a 332.5 second (roughly 5.5 minutes) cut-off has 95 % specificity and high likelihood ratios for predicting both overall cognitive and executive function impairment. This can be used as a quick and easily obtainable measure to identify people most likely to be impaired on executive and speed of processing tasks.

A few notable neuropsychological measures have used completion time to assess specific cognitive functions. For instance, Trail Making Tests (TMTs) are a set of widely accepted timed neuropsychological measures that provide insight into executive abilities.^27^ Processing speed is highly associated with performance on TMT Part B, which involves decision-making skills comparable to measures of executive function.^25,31^ Similarly, Woods et al. discovered that a patient’s question completion time on self-paced questionnaires could be used as a measure of executive functioning.^32^ Question completion time measures processing and decision-making speeds, providing insight into motivation, effort, and cognitive ability that is not measured by existing tests.^32^ These studies support the notion that timed measures may be useful as a measure of executive dysfunction in addition to their use as diagnostic screening instruments. The findings presented in our study correspond well to those reported by Woods et al. Their analysis showed that complex tasks, akin to our DOC-Cognitive tasks, were strongly related to executive function and processing speed. Their neuropsychological tests (including TMT-B and Digit Span) also correlated significantly with self-paced question completion time. Their research process was similar to ours, wherein completion time was compared to existing screens to validate completion time as a metric; both studies suggest that completion time of self-paced complex assessments may be valid markers of executive function.

Few studies use completion time of a neuropsychological screening tool as a cognitive marker. Most screens (like the MoCA or MMSE) are not routinely timed when applied in clinical settings. Moreover, executive function deficits are not often assessed in stroke patients; these deficits are subtle, challenging to test for, and often go unrecognized.^23^ Measures that are quick and easy to administer, such as DOC screen completion time, may aid in detecting executive dysfunction.

The interpretation of our findings is limited by our sample population. Compared to the total number of patients who were asked to volunteer from the stroke prevention clinic (n=1504), consenting participants (n=437) tended to be slightly younger and with slightly milder neuropsychological deficits (healthy participant bias).^12^ However, our sample also included a wide range of patients across the full spectrum of severity. As expected from stroke/TIA clinic samples, 62% had a diagnosis of stroke and/or TIA, and the rest had alternative diagnoses common in stroke prevention clinics (mimics, multiple vascular risk factors, abnormal imaging). This heterogeneity reflects the pragmatic nature of the screening and its broad generalizability to the population of patients referred to stoke prevention clinics. TIA patients are well recognized to share similar long-term risk profiles^33^ and are also at risk for cognitive impairment,^34^ compared to those with imaging confirmed strokes. The relationship between timing and gold standard testing was found across a range of severity from normal function to severely impaired. It should also be noted that there is not a single perfect cut-off score for DOC completion time that indicates executive dysfunction. To facilitate clinical utility, and because this is intended as a screen in high-volume clinics, we chose to explore a cut-off with high specificity so clinicians could be confident there was a high likelihood of true impairment beyond this time; however, this cut-off will have a low sensitivity and will miss some people with impairments. Previous work has already established that the DOC-cog score can also be a sensitive screen, effectively ruling out cognitive impairment in people who score highly.^12^ Finally, it is important to note that although screen completion time may be a useful tool to identify people likely to have executive dysfunction, it is still not equivalent to a detailed neuropsychological assessment.

## Conclusion

Clinical cognitive screening tools have not commonly used completion time as a metric. We aimed to determine whether the DOC screen completion time could provide clinically relevant information on patients’ cognitive function. DOC screen completion time reflects executive function, speed of processing and verbal fluency. When administering the DOC screen, completion time requires no additional time or patient burden to collect. This convenience is vital in busy stroke prevention clinic settings, where there is minimal time for detailed cognitive assessments. Exploring whether screen time can act as a predictor of future outcomes would provide further support the utility of this measure in clinical settings.

## Data Availability

All data is not available. DOC screening was performed as part of clinical care and used for research under REB approval with waiver of consent, on the condition of restricted data sharing.

## Non-Standard Abbreviations and Acronyms

CVLT: California Verbal Learning Test
DOC: Depression, Obstructive Sleep Apnea, Cognitive Impairment
MMSE: Mini Mental State Examination
MoCA: Montreal Cognitive Assessment
OSA: Obstructive Sleep Apnea
QCT: Question Completion Time
TMT: Trail Making Test

## Acknowledgments

We would like to acknowledge the DOC participants who volunteered their time for the study.

